# SARS-CoV-2 nucleocapsid antigen in urine of hospitalized patients with Covid-19

**DOI:** 10.1101/2021.09.28.21264239

**Authors:** N Veyrenche, A Pisoni, S Debiesse, K Bollore, AS Bedin, A Makinson, C Niel, C Alcocer-Cordellat, AM Mondain, V Le Moing, P Van de Perre, E Tuaillon

## Abstract

**Introduction:** SARS-CoV-2 nucleocapsid antigen (N-Ag) can be detected in the blood of patients with Covid-19. In this study, we used a highly sensitive and specific nucleocapsid-Ag assay to explore the presence of N-Ag in urine during the course of Covid-19, and explore its relationship with the severity of the disease.

**Material and Methods:** Urine and blood samples were collected from 82 patients with a SARS-CoV-2 infection proven by PCR and included in the COVIDotheque. We explored the presence of N-Ag in urine and blood using the AAZ N-Ag test, studied the kinetics of the marker according to the time since the onset of symptoms and evaluated the association between N-Ag levels, clinical severity and inflammation.

**Results:** In the first and second weeks of Covid-19, hospitalized patients tested positive for urinary N-Ag (81.25% and 71.79%, respectively) and blood N-Ag (93.75% and 94.87%, respectively). N-Ag levels in urine and blood were moderately correlated with the number of days after the onset of symptoms (r=-0.43, p<0.0001; r=-0.55 p<0.0001, respectively). The follow up of seven SARS-CoV-2 infected patients confirmed the waning of N-Ag in urine and blood over the course of the disease. High urinary N-Ag levels were associated with the absence of SARS-CoV-2 nucleocapsid-IgG (N-IgG), admission in intensive care units, high C-reactive protein levels, lymphopenia, eosinopenia, and high lactate dehydrogenase (LDH).

**Conclusion:** Our study demonstrate that N-Ag is present in the urine of patients hospitalized in the early phase of Covid-19. As a direct marker of SARS-CoV-2, urinary N-Ag reflects the dissemination of viral compounds in the body. Urine N-Ag is a promising marker to predict adverse evolution of SARS-CoV-2 infections.

## INTRODUCTION

The SARS-CoV-2 pandemic has upset the world and challenged medical knowledge about viral respiratory infections. One of the major characteristics of SARS-CoV-2 infection is the diversity of clinical expression, with a wide range of symptoms reported in infected persons ^1^. While severe forms constitute only a small share of Covid-19 cases, they put major pressure on health systems and are responsible for the direct excess mortality associated with the pandemic.

Although knowledge on the natural history of Covid-19 has improved considerably over the last 18 months, the exact mechanisms of impairment in the innate and adaptive immune system leading to severe forms of Covid-19 remain to be elucidated. According to our current understanding of the pathophysiology, different phases of virus and host interactions characterize SARS-CoV-2 infection ^2 3^. Following the incubation period, a high viral replication triggers the innate immune response. A strong inflammatory syndrome and the onset of the adaptive immune response characterize the second phase of Covid-19 before recovery or worsening. Severe forms of Covid-19 are associated with an excessive release of cytokines, known as a “cytokine storm”, occurring generally during the second phase of the disease ^4^. The administration of corticosteroids reduces death and time to recovery ^5^, and numerous interventional trials aim to limit this excessive inflammation using various immunomodulating treatments ^6^.

On the virus side of the host-virus interplay, the contribution to disease severity of a high SARS-CoV-2 load measured in respiratory samples remains uncertain. SARS-CoV-2 RNA detection in nasopharyngeal swabs remains the gold standard for the diagnosis of infection. The kinetics of SARS-CoV-2 RNA in the upper respiratory tract during the course of Covid-19 is well established, with high concentrations observed during the initial phase, followed by a rapid decrease in the second week after the onset of symptoms, and a low or undetectable level of RNA later ^7^. The viral loads of asymptomatic or mild forms of SARS-CoV-2 infections are similar to severe forms ^8^. Recent studies suggest that SARS-CoV-2 RNA decreases faster in mild/asymptomatic infections and in young subjects compared to severe forms and elderly subjects ^9 10^. The presence of SARS-CoV-2 components also has been reported outside the respiratory tract, such as in blood, stool and saliva. When detected, the plasma SARS-CoV-2 RNA level is generally low, and to date the virus has not been isolated from peripheral blood. Furthermore, no case of SARS-CoV-2 transmission via transfusion has been described ^11^. On the other hand, the presence of the SARS-CoV-2 nucleocapsid has been reported in the blood of Covid-19 patients. Data from two studies suggest that this circulating antigen (Ag) is detectable in almost all hospitalized patients in the early phase of the disease ^12^. A high level of nucleocapsid antigenemia (N-Ag) could be associated with severe forms of Covid-19 and the presence of circulating SARS-CoV-2 RNA ^13^. To date, the detection of antigens has only been evaluated in respiratory fluids, blood and saliva.

In this study, we used a highly sensitive and specific nucleocapsid-Ag assay to: i) explore the presence of N-Ag in the urine of hospitalized patients, ii) study the kinetics of N-Ag concentration in urine and blood during infection, iii) assess the relationship between N-Ag concentrations in urine and blood, and iv) compare urine and blood N-Ag levels in moderate versus severe forms of Covid-19.

## MATERIALS AND METHODS

### Samples and ethics

Plasma, urine and nasopharyngeal samples were collected from 82 SARS-CoV-2 infected patients admitted in Montpellier University hospitals between March 2020 and May 2021 and consenting to be included in a cohort of patients with confirmed SARS-CoV-2 (COVIDotheque cohort). A total of 82 plasma and 82 urine paired samples were taken the same day or within a maximum of 48 hours apart. In addition, series of urine and blood paired samples were collected in seven patients for a follow-up (28 paired samples). The estimated date of the onset of symptoms was recorded and ranged from 1 to 35 days before hospital admission. The SARS-CoV-2 infected patients were allocated in two groups: those hospitalized in an intensive care unit (ICU) and those not in ICU. Patient characteristics are detailed in Table 1. Controls consisted of consenting patients who were not suspected of Covid-19 and tested negative for SARS-CoV-2 RNA in nasopharyngeal samples. All tests were performed in the Virology laboratory of the CHU Montpellier. The cohort study received an institutional ethics committee approval (CPP Ile de France III, n°2020-A00935™34; ClinicalTrials. gov Identifier: <u>NCT04347850</u>).

**Table 1.**
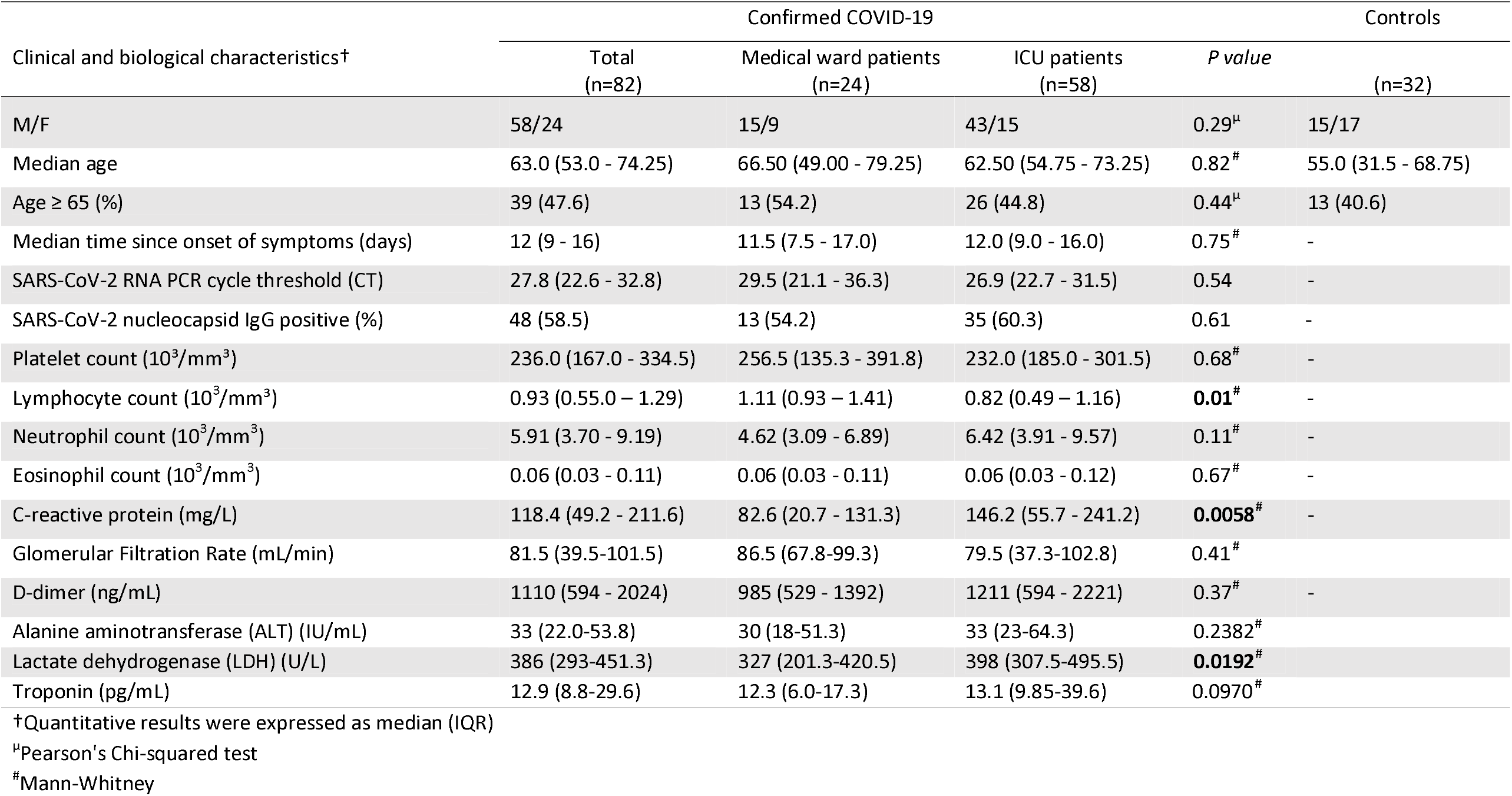
Patient characteristics

### Reverse transcription polymerase chain reaction (RT-PCR) method

RT-PCR on nasopharyngeal samples was performed by Seegene STARlet IVD for automated nucleic acids extraction and PCR setup. The Allplex 2019-nCoV Assay (Seegene) kit was used to confirm SARS-CoV-2 infection. This RT-PCR targeted SARS-CoV-2 RNA-dependent RNA polymerase (RdRP), the SARS-CoV-2 nucleocapsid (N) and the sarbecovirus envelope (E) gene. Nasopharyngeal samples were tested within a few hours after collection and without any cooling or freezing step. Various transport media were used to collect nasopharyngeal with swabs: eSwab COPAN Amies 1⍰ml, ∑-Transwab liquid Amies, viral transport medium tube VTM-M 2.0⍰ml. For each sample, the lower gene cycle threshold (Ct) was recorded.

### SARS-CoV-2 N IgG and IgA assay on plasma

SARS-CoV-N IgG and IgA detection was performed on plasma samples by indirect semi-quantitative ELISA ID Screen® ID.Vet. This assay detects IgG and IgA antibodies directed against the nucleocapsid of SARS-CoV-2 in plasma samples. The manufacturer’s instructions defined the cut-off value for a positive result: a signal ratio « Sample/Positive control% » (S/P%) ≥ 40% was considered positive, 30% < S/P% < 40% was considered suspicious and S/P% ≤ 30 was considered negative.

### SARS-CoV-2 N IgG and IgA assay on urine

SARS-CoV-N IgG and IgA detection was performed on urine samples by indirect semi-quantitative ELISA ID Screen® ID.Vet. This assay detects IgG and IgA antibodies directed against the nucleocapsid of SARS-CoV-2 in plasma samples. A signal ratio « Sample/Positive control% » (S/P%) ≥ 40% was considered positive, 30% < S/P% < 40% was considered suspicious and S/P% ≤ 30 was considered negative.

### N-antigen detection

N-SARS-CoV-2 antigen levels in urine and plasma were determined with a CE-IVD ELISA microplate assay, COVID-Quantigene® (AAZ-LMB, Boulogne-Billancourt, France). This assay detects N antigen by direct quantitative ELISA. The cut-off value was defined by the manufacturer’s instructions: samples with antigen N concentration ≥ 2.97 pg/mL were considered positive.

### Software and statistical analyses

Mann-Whitney U test was used to compare the differents quantitatives variables with non-normal distribution (Fig 1. H and I; Fig 2. A and B; Fig. 3 A and B; Fig.4 A-P). Because the distribution was non-normal and total of the discordant pairs was too low, Exact binomial’s test was used to compare the proportion of patients with urine N-Ag and/or plasma N-Ag (Fig 1.A and B). Correlations were calculated by Spearman’s coefficient (Fig 1 C-E, J and K). The median and interquartile range (IQR) were used to describe cohort characteristics. Data were analyzed using Excel 2016 (Microsoft Corp, Redmond, Washington) and GraphPad Prism 9.1.1.0 (Microsoft Corp, Redmond, Washington).

**Figure 1.**
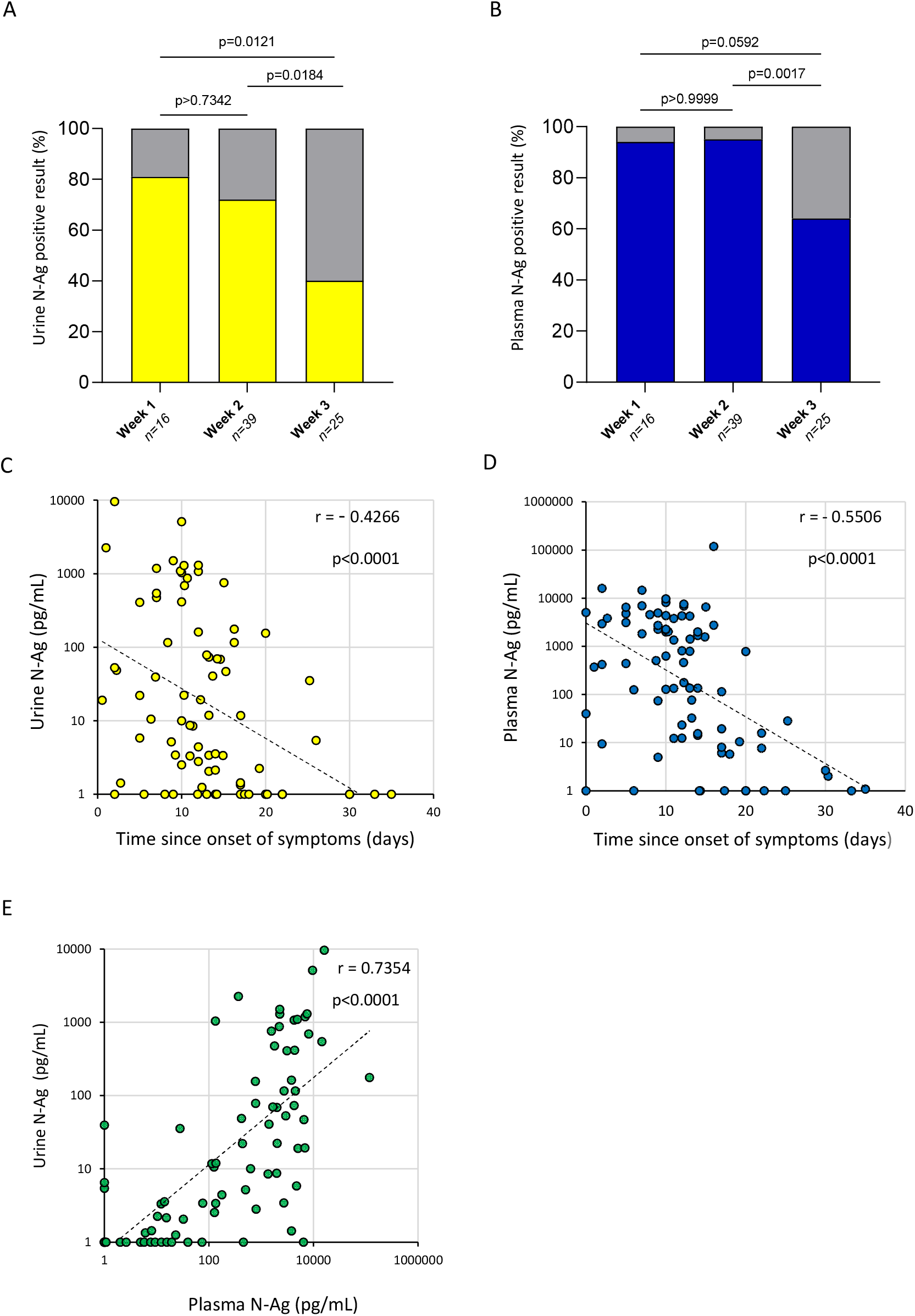
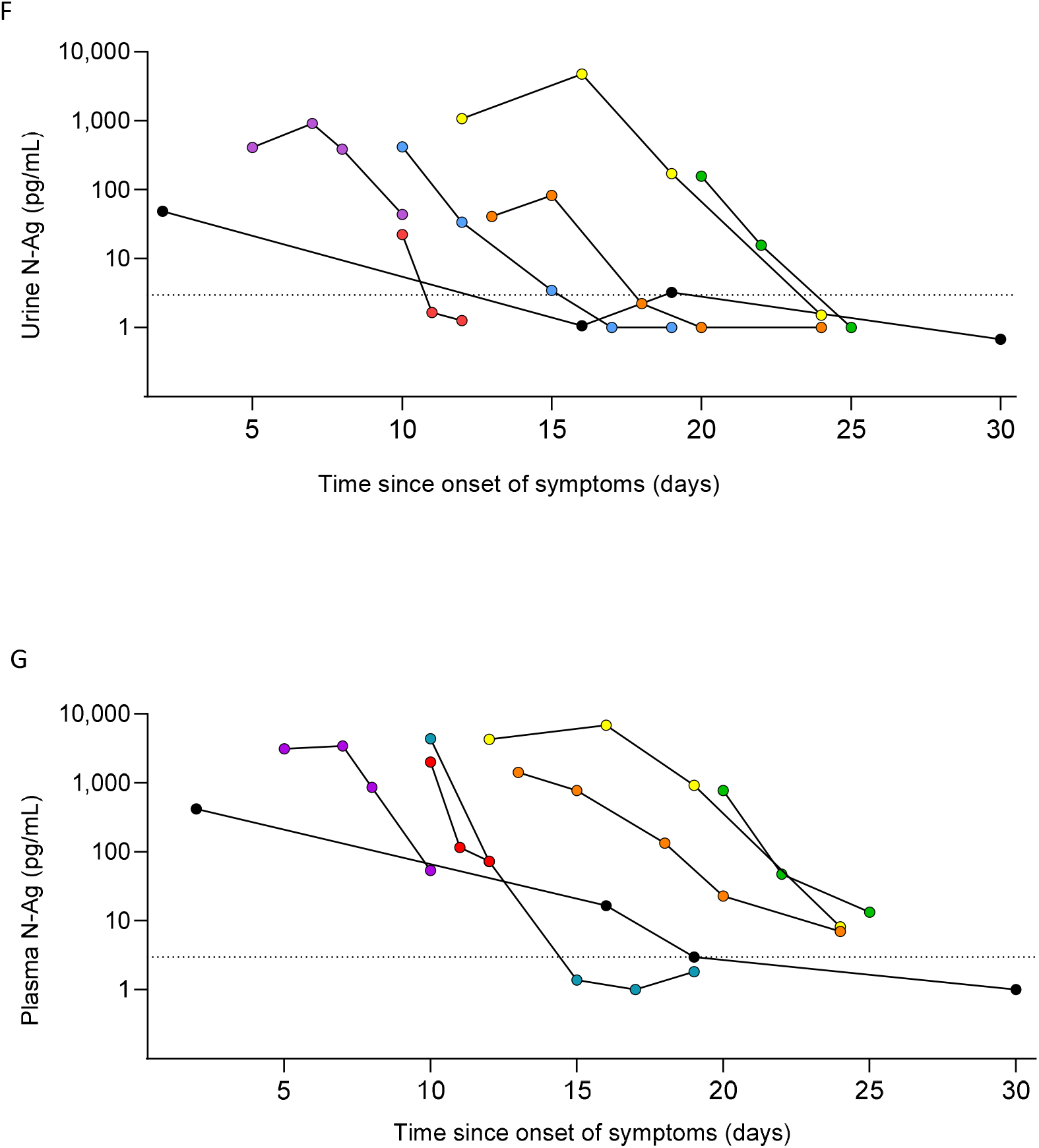
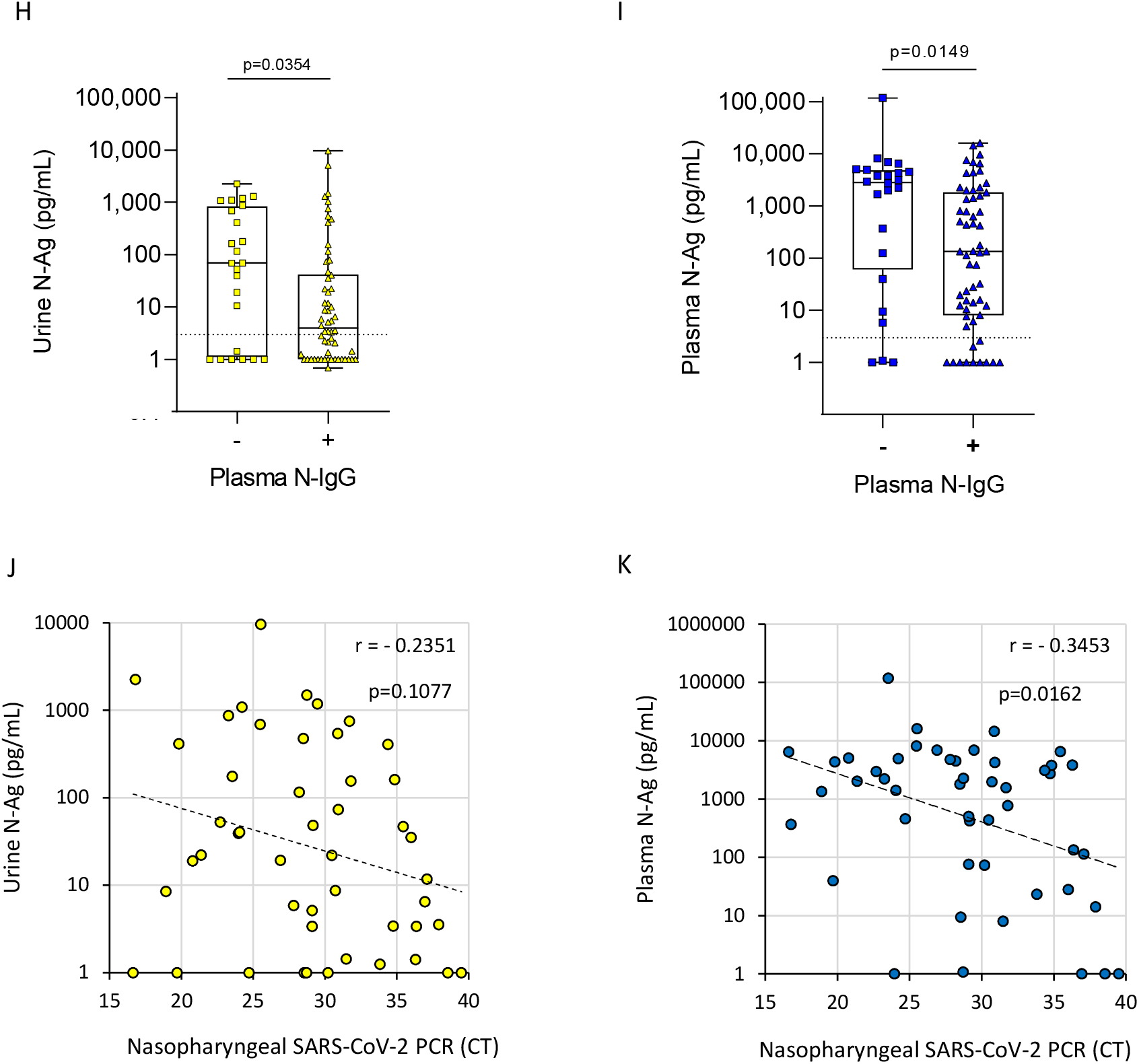
SARS-CoV-2 N-Ag in urine and plasma samples. A) Detection of urine N-Ag according to the week since the onset of symptoms in SARS-CoV-2-infected patients. Proportion of patients tested positive for N-Ag in urine were represented in yellow. B) Detection of plasma N-Ag according to the week since the onset of symptoms in SARS-CoV-2-infected patients. Proportion of patients tested positive for N-Ag in plasma were represented in blue. C) Urine N-Ag levels according to the time since the onset of symptoms, with exponential fits (dotted line). D) Plasma N-Ag levels according to the time since the onset of symptoms. E) N-Ag levels in matched urine and plasma samples. F) Longitudinal assessment of N-Ag in urine samples collected in seven patients (dotted line: lower limit of detection). G) Longitudinal assessment of N-Ag in urine samples collected in seven patients (dotted line: lower limit of detection). Matched urine and plasma samples have the same symbol and color. H) Urine N-Ag according to serological status for N-IgG. I) Plasma N-Ag according to serological status for N-IgG. J) N-Ag in urine as a function of PCR cycle threshold (Ct) in nasopharyngeal samples, with exponential fits (solid lines). K) N-Ag in plasma as a function of PCR cycle threshold (Ct) in nasopharyngeal samples, with exponential fits (solid lines).

**Figure 2.**
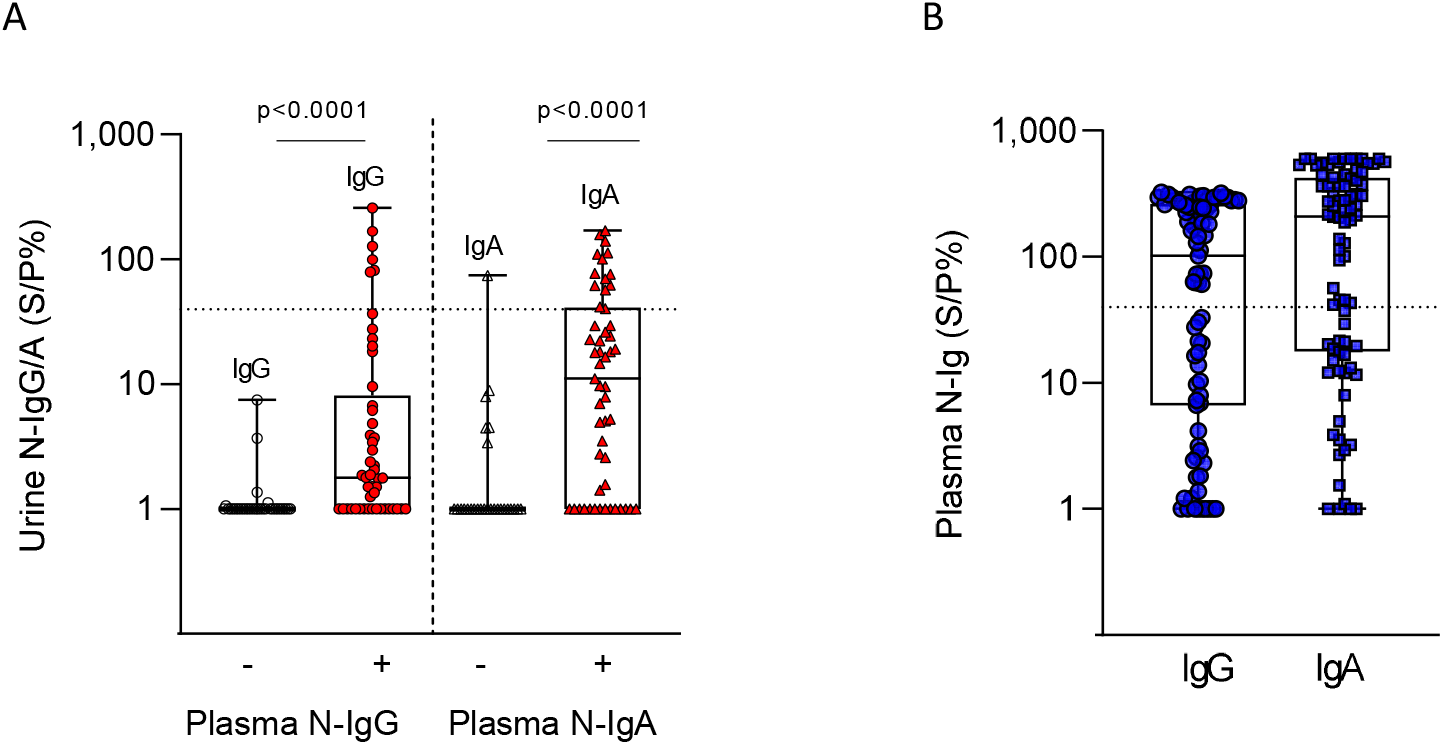
SARS-CoV-2 nucleocapsid-IgG (N-IgG) and -IgA (N-IgA) in plasma and urine samples. A) Nucleocapisd antibody levels in plasma, IgG (blue circles), IgA (blue square), limite of positivity (doted line). B) Nucleocapisd levels in urine according to plasma nucleocapsid status; urine N-IgG in plasma N-IgG negative patients (white circles) and plasma N-IgA positive patients (red circles); urine N-IgA in plasma N-IgA negative patients (white triangles) and plasma N-IgA positive patients (red triangles). Nucleocapsid antibody levels were expressed as a percentage sample/positive control ratio (S/P%).

**Figure 3.**
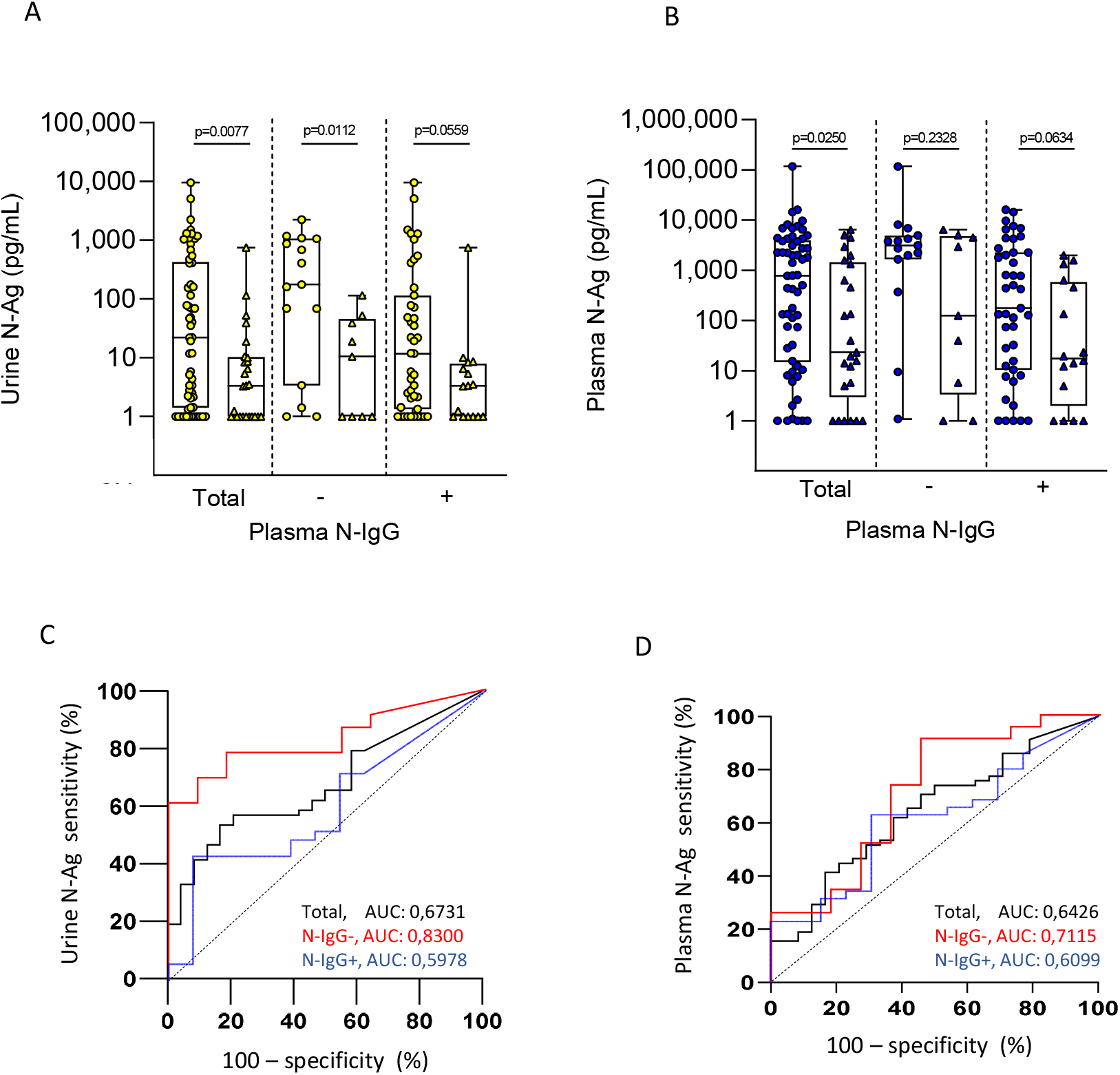
SARS-CoV-2 N-Ag according to Covid-19 severity in hospitalized patients. A) Urine N-Ag levels in patients hospitalized in intensive care units (yellow circle) versus medical wards (yellow triangle) and according to serological status for N-IgG. B) Plasma N-Ag levels in patients hospitalized in intensive care units (blue circle) versus medical wards (blue triangle) and according to serological status for N-IgG. C) Receiver operating characteristic curve (ROC) evaluating the ability of urine N-Ag levels to discriminate patients hospitalized in intensive care units versus medical wards (Black: all patients; red: nucleocapside-IgG seronegative patients; blue: nucleocapsid-IgG seropositive patients). D) ROC evaluating the ability of plasma N-Ag levels to discriminate patients hospitalized in intensive care units versus medical wards (Black: all patients; red: nucleocapside-IgG seronegative patients; blue: nucleocapsid-IgG seropositive patients).

**Figure 4.**
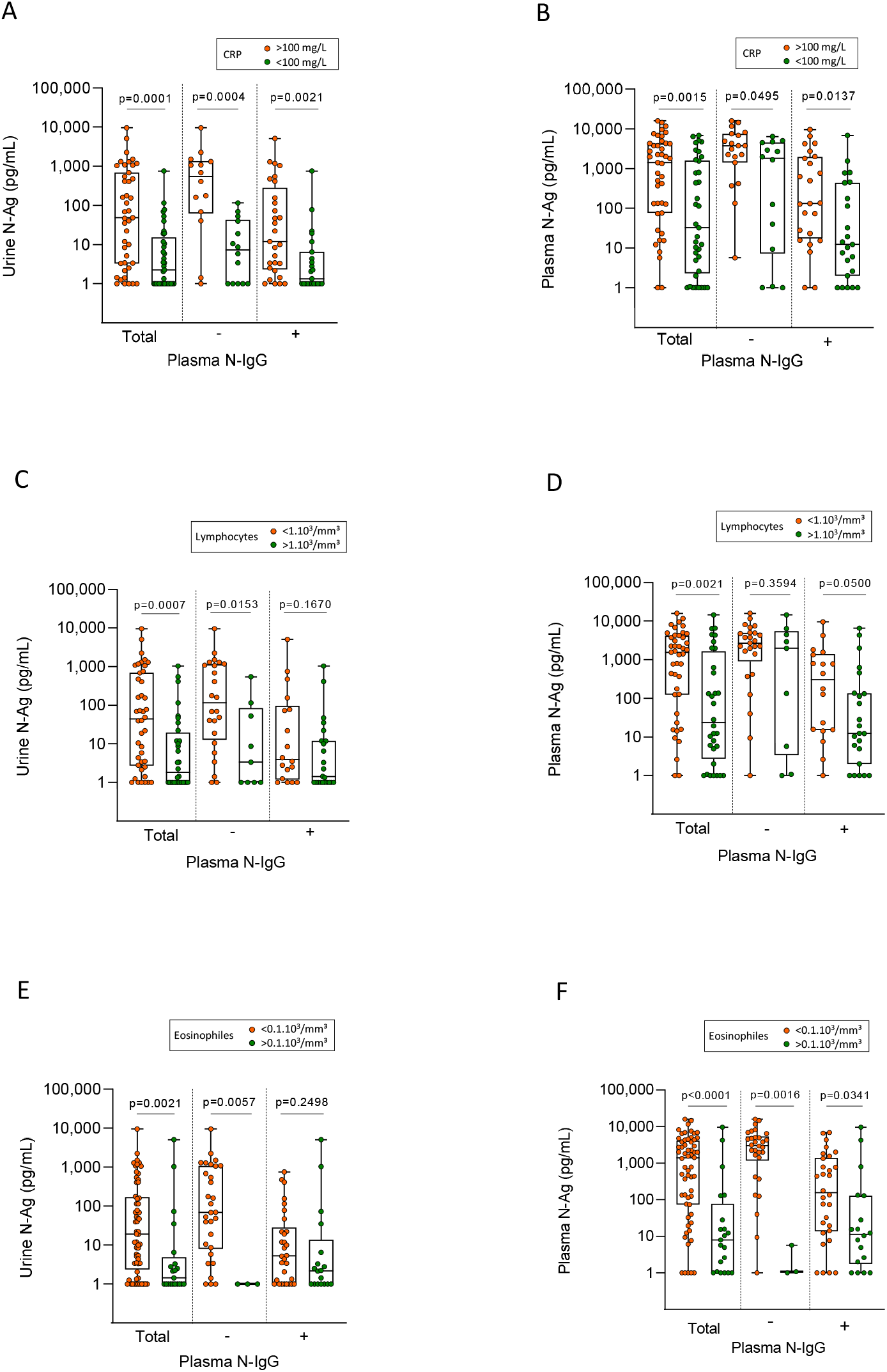
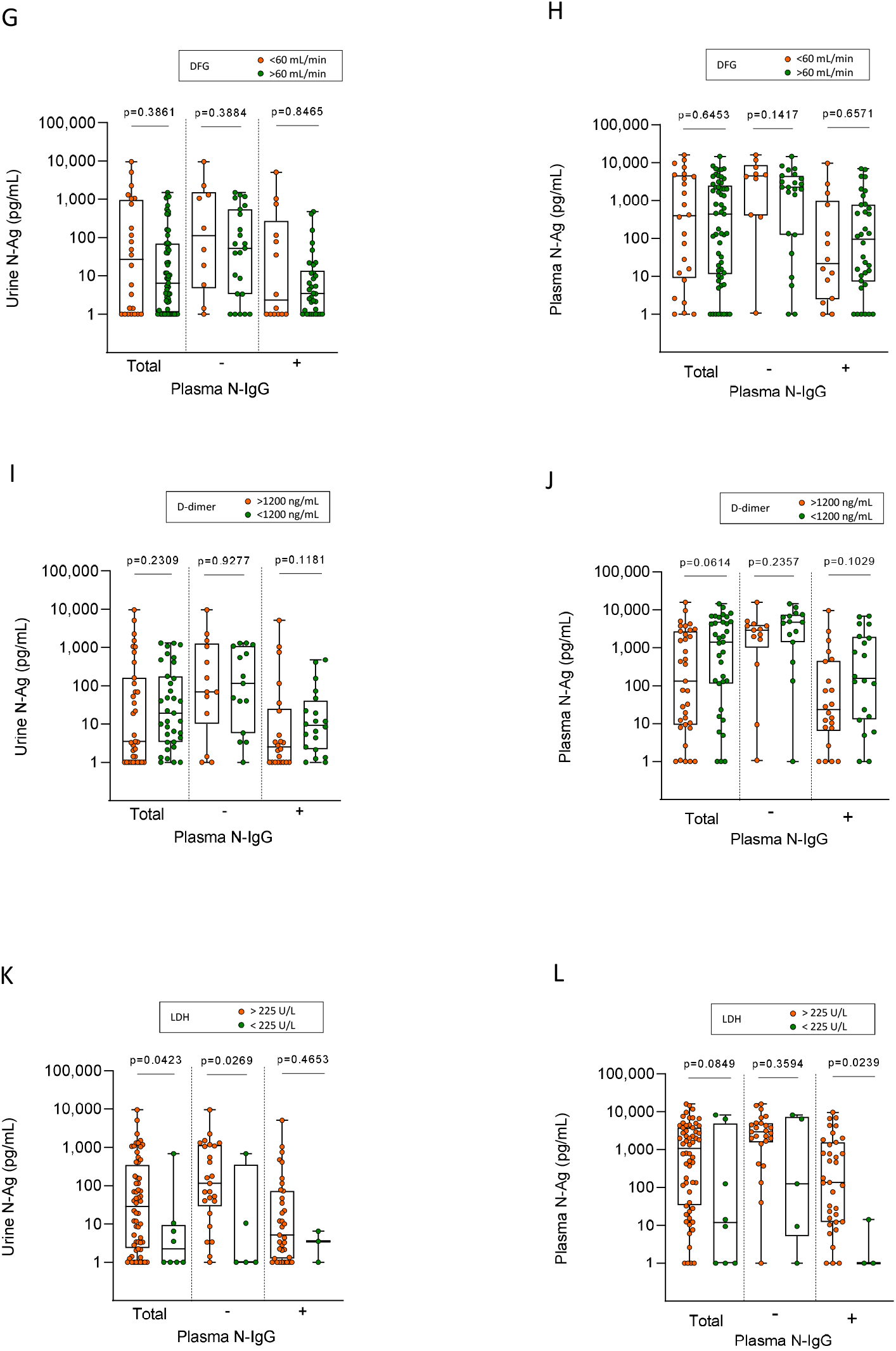
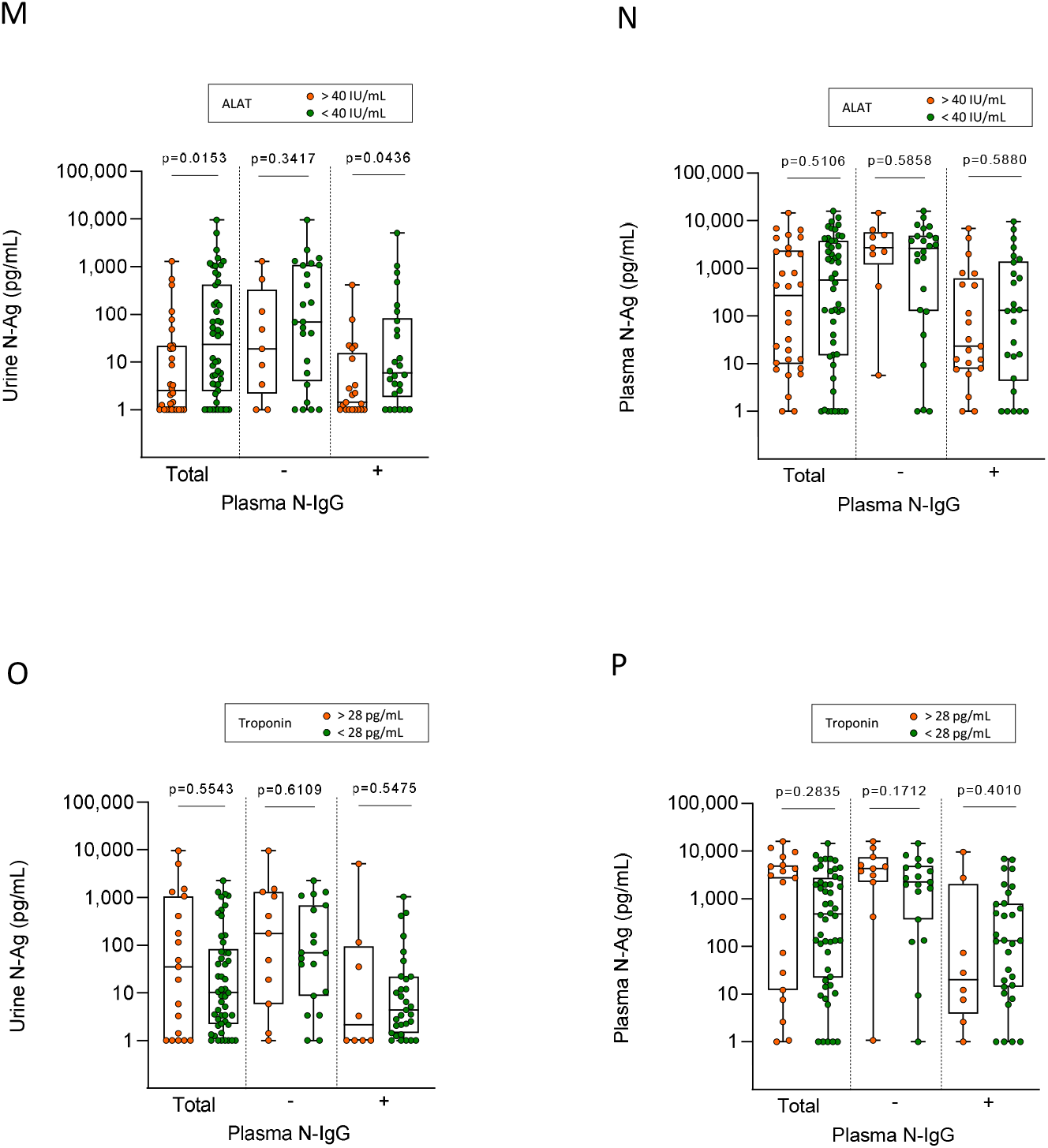
Association of N-Ag levels with biological indicators of Covid-19 severity. A) N-Ag levels in urine according to C-reactive protein (CRP) and N-IgG serological status. B) N-Ag levels in plasma according to CRP and N-IgG serological status. C) N-Ag levels in urine according to lymphocyte count and N-IgG serological status. D) N-Ag levels in plasma according to lymphocyte count and N-IgG serological status. E) N-Ag levels in urine according to eosinophil count and N-IgG serological status. F) N-Ag levels in urine according to eosinophil count and N-IgG serological status. G) N-Ag levels in urine according to Glomerular Filtration Rate (GFR) and N-IgG serological status. H) N-Ag levels in plasma according to GFR and N-IgG serological status. I) N-Ag levels in urine according to D-dimer level and N-IgG serological status. J) N-Ag levels in urine according to D-dimer level and N-IgG serological status. K) N-Ag levels in urine according to lactate dehydrogenase (LDH) level and N-IgG serological status. L) N-Ag levels in plasma according to LDH levels and N-IgG serological status. M) N-Ag levels in urine according to alanine aminotransferase (ALT) level and N-IgG serological status. N) N-Ag levels in plasma according LDH level and N-IgG serological status.

## RESULTS

### N-Ag is present in the urine of patients hospitalized with SARS-CoV-2 infection

We tested the presence of N-Ag in urine samples of 82 patients hospitalized for a SARS-CoV-2 infection confirmed by RT-PCR. N-Ag was present in the urine of 41/55 patients (74.55%) tested during the first two weeks after the onset of symptoms, (Se: 81.25% and 71.79%, respectively) with concentrations ranging from 1.00 to 9613.03 pg/mLFig. 1.A). All urine samples from control subjects tested negative for N-Ag (Sp: 100%; data not shown). In blood-paired samples, N-Ag was detected from 53/55 (94.64%) patients tested during the first two weeks after the onset of symptoms (Se: 93.75% and 94.87%, respectively) with concentrations ranging from 1.00 to 16019.30 pg/mL (Fig. 1.B). Urine N-Ag levels correlated with blood concentrations (r=0.7354) (Fig. 1.E).

### Urinary N-Ag concentrations decrease over time but remain detectable during the second week after the onset of symptoms and after nucleocapsid IgG (N-IgG) seroconversion

N-Ag levels in urine was inversely correlated with the number of days after the onset of symptoms (fig. 1.C) (r = −0.43; p<0.0001). Urinary N-Ag levels were high in samples collected during the first and second week after the onset of symptoms and decreased sharply during the third week (Supplemental Figure S1.A). Blood N-Ag correlated well with the number of days after the onset of symptoms (r = −0.55; p<0.0001), and also decreased after the first two weeks (Supplemental Figure S1.B). The follow-up of seven SARS-CoV-2 infected patients confirmed the gradual decrease of urine and blood N-Ag levels over time (Fig 1.F, G).

The analysis of blood N-Ag according to serological status against SARS-CoV-2 nucleocapsid showed lower concentrations in N-IgG positive patients compared to N-IgG negative patients. However, most patients (81.36%) who tested positive for SARS-CoV-2 nucleocapsid IgG remained positive for blood N-Ag (Fig 1.I). The level of N-Ag in the urine was also significantly lower in patients who tested positive for circulating anti-N IgG compared to N IgG negative patients, but we observed a less pronounced difference between the two groups than in blood (Fig 1.H-J. CT results of SARS-CoV-2 RNA PCR in nasopharyngeal swabs were not correlated with urine N-Ag level (r=-0.23, p = 0.1077; Fig 1.K), and weakly correlated with blood N-Ag (r=-0.34, p = 0.0162; Fig 1.K).

Urine samples also were assessed for antibodies against SARS-CoV-2 nucleocapsid (Figure 2). Patients tested negative for nucleocapsid antibodies in plasma tested also negative for both N-IgA and N-IgG in urine. Signal ratios for N-IgA and N-IgG were higher in urine samples collected after seroconversion has occurred (p <0.0001). Using the same threshold as for plasma, N-IgA were detected in 17/82 patients (20.7%), and N-IgG in 6/82 (7.3%) patients. All urine samples from SARS-CoV-2 negative controls tested negative for N-IgG and N-IgA (data not shown).

### High urinary N-Ag levels are associated with admission in intensive care units

Patients had higher urinary N-Ag levels when hospitalized in intensive care units (ICU) compared to medical wards (p=0.0077) (Fig. 3.A). The levels were higher in ICU seronegative hospitalized patients (p=0.0112), and a trend toward a higher level of N-Ag was observed in ICU seropositive patients (p=0.0559) (Fig 3.A). Blood N-Ag levels were also higher in patients hospitalized in ICU compared to medical wards (p=0.0250) (Fig 3B). The ROC curves show that both high urine and blood N-Ag levels were predictive of a condition requiring intensive care (Fig 3.C, 3.D). A higher accuracy was observed for urine N-Ag as a predictor of severe Covid-19 compared to blood N-Ag, especially on seronegative patients (AUC: 0.8300 vs 0.7115, respectively).

### Relationship of urine and blood N-Ag levels with biological markers of Covid-19 severity

Levels of urine and plasma N-Ag were analyzed according to abnormalities in biological markers associated with Covid-19 severity (Fig 4.). Higher urine and blood N-Ag levels were measured among patients with a C-reactive protein (CRP) over 100 mg/L. The differences remained significant when N-Ag levels in urine and blood were analyzed according to N-IgG serological status. Higher urine and blood N-Ag levels also were measured among patients with lymphopenia and low eosinophil counts. Urine N-Ag levels were higher when the Lactate dehydrogenase (LDH) level was elevated. We did not observe an association between N-Ag levels and low platelet counts (<200/µL), abnormal troponin levels (>60 mg/mL), high D-dimer levels (> 1200 mg/mL), or the glomerular Filtration Rate (GFR) based on the Modification of Diet in Renal Disease (MDRD) study equation (< 60 ml/min/1.73m2). Urine N-Ag levels were lower in patients with elevated alanine aminotransferase (ALT) concentrations.

## DISCUSSION

In this study, we assess N-Ag in urine of patients hospitalized for a Covid-19 infection confirmed by PCR. We analysed N-Ag levels in urine according to time after symptoms onset, level of antigenemia, serological status and severity. Our results demonstrate that N-Ag is present in urine of patients hospitalized for a SARS-CoV-2 infection. Urinary N-Ag concentrations decreased progressively after the onset of symptoms following two phases: a low decay during the first and second weeks, and a sharp decrease during the third week. The presence of circulating antibodies against nucleocapsid was associated with a lower level of urine N-Ag, but the SARS-CoV-2 nucleocapsid remained detectable in the urine samples of most of the patients seropositive for N-IgG.

It is thought that severe forms of the disease are caused by an inappropriate host response to SARS-CoV-2, with an inefficient innate response during the first week after the onset of symptoms, followed by an over inflammation corresponding to systemic cytokine storm thereafter. Steroid treatments reduce mortality related to Covid-19 in hospitalized patients requiring oxygen ^14^. Studies using interleukin-6 antagonist Tocilizumab also have obtained encouraging results ^15 16^. Levels of SARS-CoV-2 RNA in the upper respiratory track do not appear to be a reliable marker of Covid-19 severity since high concentrations of virus in nasopharyngeal and saliva specimens are observed in asymptomatic, mild and severe forms of SARS-CoV-2 infections ^17^. Blood SARS-CoV-2 RNA is more frequently detectable and found at higher levels in severe forms of Covid-19 ^18 19^. SARS-CoV-2 viremia is associated with disease severity, patient outcome and inflammatory biomarkers ^20^. Ciliated and AT2 cells in airway and alveolar regions are the first targets of SARS-CoV-2. However, systemic clinical manifestations suggest that SARS-CoV-2 also can infect different organs through the bloodstream, such as endothelial cells ^21^, gastrointestinal cells and ACE2 positive distal tubule cells ^22^. Furthermore, the administration of convalescent plasma therapy ^23^ and monoclonal antibodies (mAb) against the Spike protein help improve Covid-19 recovery ^24^. All together, these observations suggest that SARS-CoV-2 replication and plasma viremia may contribute severity of Covid-19.

The first reports on Covid-19 infrequently detected circulating SARS-CoV-2 RNA ^25^. Recent studies have inconsistently detected SARS-CoV-2 RNA, and generally with a low viral load ^26,27^. In contrast, in the study of Le Hingrat et al. using the same assay as in our study, N-antigenemia appeared to be a sensitive marker of SARS-CoV-2 infection in hospitalized patients, able to provide a surrogate test to molecular approaches ^12^. Dandan S. et al. confirmed this observation using a digital enzyme-linked immunosorbent method ^13^. In line with these studies, we observed that the N-Ag levels in our population of hospitalized patients most of the time were over 100 pg/mL during the first week after the onset of symptoms, and remained largely over the lower limit of quantification of the assay during the second week. Given the much better analytical sensitivity of PCR methods compared to antigen immunoassays, these findings are surprising. SARS-CoV-2 nucleocapsid antigen may be released in the bloodstream or circulate after destruction of the virus particle and virus RNA. Additionally, SARS-CoV-2 nucleocapsid antigen produced in excess during profuse viral replication and non-included in virions.

Detection of SARS-CoV-2 RNA in urine specimens has been reported in less than 5% of confirmed Covid-19 cases ^28^. Using mass spectrometry, Mishra C et al. have reported detection of nucleocapsid-derived peptides in urine of a third of Covid-19 patients ^29^. Our results using a highly sensitive immunoassay demonstrate the presence of N-antigen in urine in most of Covid-19 hospitalized patients during the first two weeks after onset of the symptoms.

The kidney is among the most frequently affected extrapulmonary organs during SARS-CoV-2 infection, and varying degrees of renal damage have been reported in COVID-19 patients ^30,31^. Acute kidney disease was observed in a quarter of the patients included in our study. We observed high concentrations of urine N-Ag during the first two weeks after the onset of symptoms, and no association between urine N-Ag levels and altered renal function. The origin of N-antigen detected in urine is uncertain. N-Ag in urine may originate from the blood and be excreted by the kidney, as suggested by the correlation between N-Ag concentrations in blood and urine. The SARS-CoV-2 nucleocapsid is a 46 kd protein. The glomerular permeability and filtration probably permit the excretion of this small-size protein. However, the C-terminal region of the SARS-CoV-2 nucleocapsid possesses binding affinity to form dimers that produce large compounds that may be not be filtered by the kidney. Furthermore, after seroconversion against the nucleocapsid, immune complexes also form, with a size that makes them unable to be filtered. Of note, although Covid-19 associated glomerular disease has been reported, this type of kidney injury seems infrequent among acute kidney diseases associated to SARS-CoV-2 infection ^32^. A local production of SARS-CoV-2 nucleocapsid may be another possible origin of the antigen detected in urine. Alongside hypoxia, circulating disorder and inflammation, SARS-CoV-2 infection may directly contribute to kidney injury. Cells expressing ACE2 are present in the tubules, and studies have shown that SARS-CoV-2 RNA, nucleocapsid and spike protein accumulate in tubules ^33 22^.

Both blood and urine N-Ag levels may reflect SARS-CoV-2 disseminated infection. We observed a moderate correlation between N-Ag in blood and urine, but the kinetic of this marker may be different in these two compartments. The development of anti-nucleocapsid humoral response may induce the formation of immune complexes that interfere with N-Ag quantification in blood. Hence, after seroconversion N-Ag and N-IgG levels may represent only the unbound fraction available to be measured by the immunoassays. In urine, antibodies directed against nucleocapsid were only detected in one quarter of the patients who tested positive for circulating N-IgG, limiting the risk of underestimation of N-Ag levels by the immunoassay. In other words, N-Ag levels in urine may be more accurate since nucleocapsid Ag quantitation in urine is probably less impacted by the formation of immune Ag-Abs complexes compared to blood. N-Ag in urine may better reflect disseminated infection than nucleocapsid antigenemia, especially after seroconversion against SARS-CoV-2 nucleoprotein. Besides interfering with assay measurement, the presence of circulating antibody-antigen complexes may bind FC receptors activating monocytes/macrophages, and fuel the hyperinflammation observed in the second phase of Covid-19 ^34^.

Risk factors related to age and comorbidities, alongside inflammation and cytopenia, are associated with the development of severe forms of Covid-19 requiring hospitalisation and intensive care. At present, however, the progression to a severe form of Covid-19 remains unpredictable. We observed higher concentrations of urine N-Ag in samples collected in patients hospitalised in intensive care units compared to medical wards. This result is in line with the study of Caceres P. et al., reporting that SARS-CoV-2 viral load in urine sediments was associated with higher mortality in hospitalized patients ^17^. Furthermore, in our study, urine and plasma N-Ag levels were associated with several early markers of Covid-19 severity, such as lymphopenia, low eosinophil count, and CRP levels. We observed lower levels of urine N-Ag in patients with abnormal ALT, which may be because liver injury is delayed after the first week in the course of prolonged Covid-19 while the decay of N-Ag is already underway ^35^.

Our study has several limitations. The population is not representative of SARS-CoV-2 infected individuals. All of the subjects had mild or severe forms of Covid-19 and required oxygen, whereas a majority of SARS-CoV-2 infected individuals do not require hospitalization. In addition, patients requiring critical care are over-represented because their urine samples frequently were collected in routine care. We did not assess the value of N-Ag as a predictive marker of adverse evolution but only as a marker associated to severe Covid-19. Finally, N-Ag levels were analysed on a single urine sample while results on urine samples collected taken over a 24-hour period would be more accurate.

In conclusion, these results demonstrate that N-Ag is present in urine of patients hospitalized for Covid-19. As a direct marker of SARS-CoV-2 infection, urinary and blood N-Ag reflect the dissemination of viral compounds in the body and probably SARS-CoV-2 replication. Further studies are required to evaluate the value of urinary N-Ag to predict the adverse evolution of SARS-CoV-2 infections.

## Supporting information

SFig.1

## Data Availability

The datasets analysed during the current study are available from the corresponding author on reasonable request.

## ACKNOWLEDGMENTS

The author would like to thank Grace Delobel for English language editing and review services. This work was funded by the Montpellier University Hospital, Muse I-SITE Program Grant, University of Montpellier.

## AUTHOR CONTRIBUTIONS

N. V. has performed experiments, statistical analysis, and wrote the manuscript. A. P., C. N. and C. A. C. have performed experiments. A. P. has helped to use GraphPad Prism 9.1.1.0. S. D., K. B., A.-S. B., A.M. M., A. M., V. L. M. and P. V. P. have discussed the results and critically reviewed the manuscript. E. T. has conceived the study, discussed the results and wrote the manuscript. All authors contributed to the article and approved the submitted version.

## DATA AVAILABILITY STATEMENT

The data that support the findings of this study are available from the corresponding author upon reasonable request.

