## Supplementary material for "SARS-CoV-2 nucleocapsid antigen in urine of hospitalized patients with Covid-19": SFig.1

### Supplemental Figure S1.A

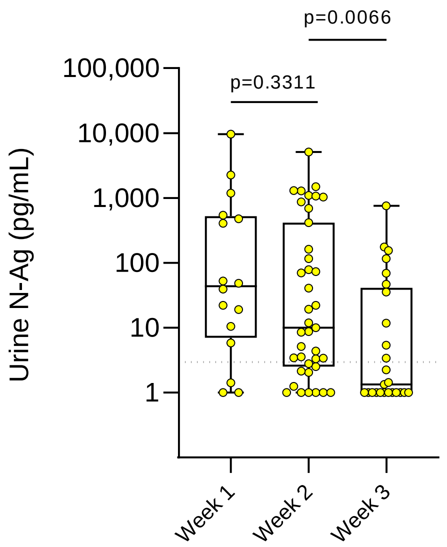


### Supplemental Figure S1.B

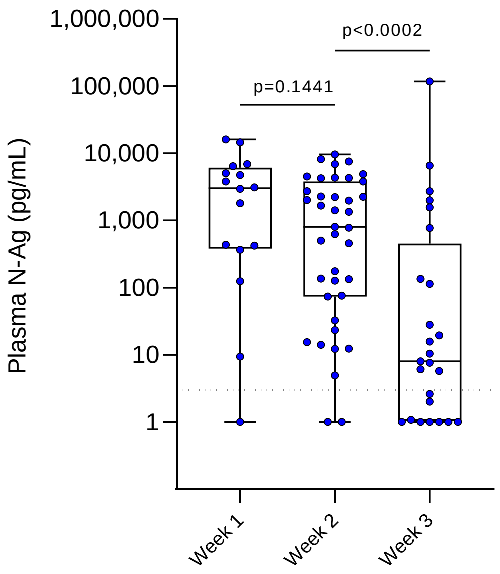


**Supplemental Figure S1.A.** Urine N-Ag levels according to the week since the onset of symptoms in SARS-CoV-2-infected patients

**Supplemental Figure S1.B.** Plasma N-Ag levels according to the week since the onset of symptoms in SARS-CoV-2-infected patients
